# Small fibre neuropathy detected by ultrasound of median nerve in children with type 1 diabetes mellitus is related to glycemic control

**DOI:** 10.1101/2025.05.24.25328284

**Authors:** Miriam C. Eilers, Metsnanat Fellmann, Dagmar l’Allemand, Janina Wurster, Sandro Meier, Jürg Lütschg, Marc Grüner, Erin West, Ute Muhitira, Sarah S. Oberhauser, Katrin Heldt, Philip J. Broser

## Abstract

**Aim:** The initial stage of diabetic peripheral neuropathy (DPN) is characterized by the damage of small nerve fibres. High-resolution ultrasound is used to assess the nerve cross-sectional area (CSA), representing these. The aim of our study was to study the influence of metabolic control as measured by HbA1c and body surface area (BSA) on CSA.

**Method:** 73 children with type 1 diabetes mellitus (T1D) (8-16 years) and 71 healthy controls 8-16 years) underwent ultrasound of the median nerve. The association of CSA with BSA and HbA1c was tested with linear regression models.

**Results:** In children with HbA1c >9%, CSA was significantly higher than in controls. In the combined group including all children, CSA was significantly correlated with BSA. In children with T1D, linear models taking these effects of BSA into account showed a highly significant correlation of the CSA with the HbA1c (Regression Coefficient 0.217 (CI 0.073 - 0.361)).

**Conclusion:** Median nerve CSA is related to BSA and HbA1c. When CSA of the median nerve is adjusted for BSA a clear and robust correlation with thicker nerves with higher HbA1c was found.

Based on this data, we derived a method for the early prediction of small fibre neuropathy in children with T1D.

**Highlights:**

- Ultrasound measurement of median nerve thickness is a non-invasive diagnostic marker that can be used for early detection of nerve damage.
- Children with type 1 diabetes have a thicker median nerve than healthy controls.
- Nerve thickening depends on metabolic control: the worse the HbA1c, the thicker the nerve.
- Nerve thickness is measured as the diameter of the median nerve (cross-sectional area, CSA). It represents the damage to the small unmyelinated fibres that are affected in the early stages of diabetic neuropathy (DNP).

## 1. Introduction

Diabetic peripheral neuropathy (DPN) is a serious, long-term microvascular complication of T1D that affects small unmyelinated nerve fibers first. The cardinal symptoms of DPN, such as pain and changes in temperature sensation, are rare in childhood and adolescence; however, subclinical pathology may be present early after the onset of T1D [1,2]. Screening for small fiber function is recommended [3], but the assessment of temperature and pinprick sensation is highly dependent on clinician experience and patient compliance. In children, this contributes to the challenge of detecting DPN [4], which may also explain the high variability in prevalence described in this age group (3–19%) [5,6].

The current gold standard for the diagnosis of DPN is the measurement of nerve conduction velocity (NCV) [7]. Reduced NCV in children is a sign of neuropathy [8,9]. As shown by Oberhauser et al., NCV starts to decline as early as at the end of the remission phase [10]. Although objective, quantifiable, and sensitive, the NCV test only detects changes in large myelinated fibers, which is a major limitation. In all nerves studied, sensory axons, which also represent small fibers, outnumber motor axons by a ratio of at least 9:1 [11]. Therefore, NCV remains normal at the onset of neuropathies, although small unmyelinated nerve fibres are damaged early on [12]. High-resolution nerve ultrasonography is a non-invasive tool for assessing the structure of peripheral nerves [13]. It allows for the recording of the nerve cross-sectional area (CSA) and there also for the assessment of the structure of A-delta and C-nerve fibers, which are predominantly thin and unmyelinated (also known as small fibres or sensory fibers). In adults with diabetes, studies have shown CSA enlargement of the median nerve and the posterior tibial nerve [14]. However, there is a lack of studies on the use of nerve ultrasound in children, including normative data. Thus, the primary objective of our study is to investigate whether the CSA is enlarged in children with T1D and whether a correlation with the quality of glycemic control exists.

It is important to know that the CSA of the median nerve is strongly correlated to the Body Surface Area [15]. In neuropediatrics, BSA, as calculated based on Du Bois formula, is a well-established number that reflects the size of the body area a of the skin and so the size of the receptive field of the peripheral nervous system.

HbA1c level is a well-established and recommended marker for monitoring glycemic control [16]; the target for patients with T1D is ≤7%. An increased level is a significant risk factor for vascular complications [17] and was also found to be an important risk factor for the development of subclinical neuropathy [2,18,19]. We hypothesize that good glyemic control (HbA1c ≤7%) will prevent or delay the onset of neuropathic complications and small-fibre neuropathy at an early stage of the disease, whereas suboptimal glycemic control would gradually lead to neuropathic complications We therefore divided HbA1c levels between good (HbA1c ≤7%), moderately insufficient (≥7-9%) and insufficient (HbA1c ≥9%).

Based on these assumption we built a model for the early detection of structural changes of the peripheral nerve system, which also includes changes to the unmyelinated fibers [20-24].

## 2. Research Design and Methods

This cross-sectional retrospective study was conducted between 2022 and 2023. The study was approved by the Ethics Committee of Eastern Switzerland (approval no.: 2022-00216). Written informed consent was obtained from the participants’ caregivers before the start of the study along with additional self-reported consent for children > 12 years.

73 children aged > 8 and < 16 years diagnosed with T1D at least 6 months prior were eligible for inclusion in the study group. They were seen annually at the neurophysiological department of our hospital. The definition of diabetes as used in this study was based on the International Society for Pediatric and Adolescent Diabetes (ISPAD) Clinical Practice Consensus Guidelines [25]. T1D is defined as beta-cell destruction, usually leading to absolute insulin deficiency. It is immune-mediated and characterized by the presence of one or more autoimmune markers (GAD, IA2, IAA, ZnT8, pancreatic cell antibody) or idiopathic. Children with chronic diseases other than T1D (i.e., hereditary and inflammatory diseases) were excluded.

Children with mild traumatic nerve injuries are frequently cared for at the local children’s hospital. These children typically have one unaffected body side and were therefore eligible as controls. Using the same exclusion criteria as in the diabetic group, 71 children were included in the control group.

Using high-frequency (>18 MHz) ultrasound probes with an axial spatial resolution of 50 µm, it is possible to detect even small and subtle changes to the CSA of the median nerve. Ultrasound examinations were performed using a Canon Aplio i800 (Canon Medical Systems, Otawara, Japan) ultrasound scanning machine. An i22LH8 probe with a scanning frequency of 22 MHz was used. For the ultrasound examination of the median nerve, we used the formal labelling standardized by Jenny et al. (25).

The ultrasound examinator was blinded to the laboratory results (HbA1c). Location R (L) 1 represents the right (left) median nerve proximal to the flexor retinaculum. Location R (L) 2 represents the right (left) median nerve in the middle of the forearm, where the nerve is loosely embedded between the superficial and deep finger flexors (Figure 1A, B). Previous studies had shown that there is no difference between left and right side [26], therefore the data from both sides were taken. If both sides could be measured, the mean of both sides was used, otherwise the data from the measured side was used. During the scan, the forearm was fully supinated. The transducer was positioned at a 90° angle to the surface, and sufficient gel was applied [23]. The median nerve can be recognized in the ultrasound image by its honeycomb pattern (Figure 1C, D).

**Fig. 1:**
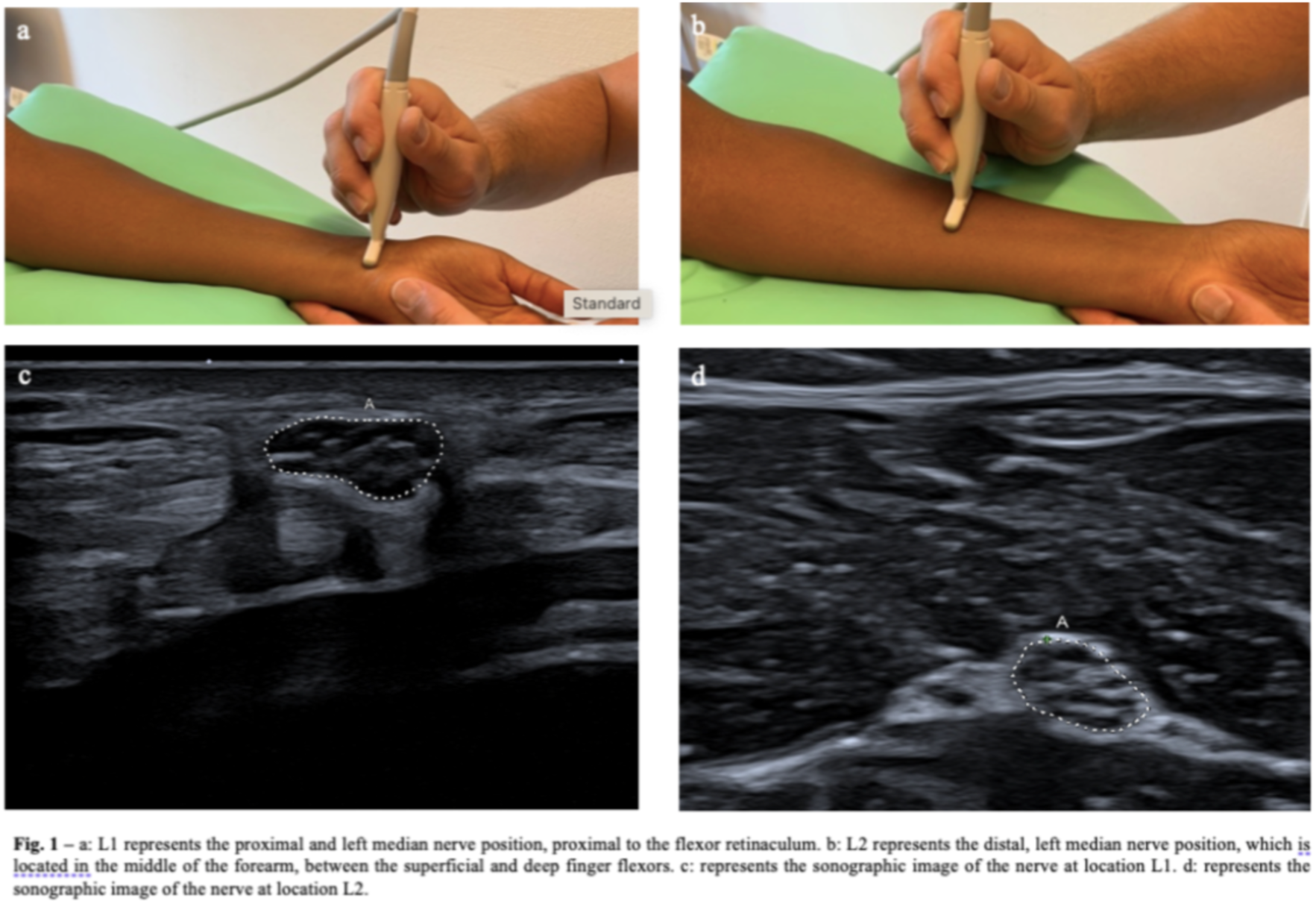
Ultrasound sensor position at location 1 (L1) and Location 2 (L2).

The outline of the nerve was drawn using a freehand tracing tool, and the border between the nerve and connective tissue was imaged. The tool then calculated the CSA and perimeter [26].

## 3. Data Processing and Statistical Computation

After the measurement procedure the data was stored in our custom made KIND database, for further processing, together with height, weight and HbA1c.

With a significance criterion of α = 0.5 and 0.8 power, the minimum sample size needed to detect small differences in CSA between the diabetes and control groups according to Velleman and Welsch [27] was n = 20. All analysis was performed using the R statistical program [28]. Two-sample Wilcoxon tests (Mann–Whitney test) or Kendall’s tau were used to perform between-group comparisons. Correlation between BSA and CSA was calculated using Pearson correlation test. Linear models relating CSA with HbA1c and BSA were built. The multiple linear regression model was fitted and tested for significance by a two-way ANOVA Type 3 [29].

## 4. Results

The characteristics of the 73 children and adolescents with T1D (diabetes group) and 71 children without T1D (control group) are shown in Tab. 1.

**Table 1:** Group characteristics.

| Table 1 |  |  |  |  |
| --- | --- | --- | --- | --- |
| Characteristic | Healthy N = 71<sup>1</sup> | Low_HbA1c N = 25<sup>1</sup> | Mid_HbA1c N = 35<sup>1</sup> | High_HbA1c N = 13<sup>1</sup> |
| Age [y] | 12.42 (9.83, 14.22) | 12.83 (11.52, 14.72) | 12.68 (10.43, 14.12) | 14.08 (12.96, 14.99) |
| Sex |  |  |  |  |
| m | 23 (32%) | 13 (52%) | 16 (46%) | 7 (54%) |
| w | 48 (68%) | 12 (48%) | 19 (54%) | 6 (46%) |
| Body height [cm] | 159 (147, 164) | 161 (149, 172) | 154 (145, 164) | 164 (153, 171) |
| Percentile Length | 69 (54, 91) | 79 (50, 91) | 74 (34, 86) | 69 (54, 90) |
| Weight [kg] | 50 (38, 58) | 53 (43, 64) | 48 (40, 60) | 50 (42, 62) |
| Percentile Weight | 64 (45, 92) | 73 (49, 92) | 72 (43, 88) | 53 (19, 86) |
| HbA1c |  | 6.60 (6.40, 6.90) | 7.80 (7.40, 8.10) | 12.90 (10.10, 14.50) |
| <sup>1</sup> Median (Q1, Q3); n (%) |  |  |  |  |

CSA was measured at location 1 and 2. At location 1 no difference in CSA was found between healthy children and children with T1D (Fig. 2 Panel A). This was different at location 2, where the nerve is only loosely embedded between the superficial and deep flexor muscles. Here, an increase in CSA could be seen, however this was only significant for the group with HbA1c >9% (Fig. 2, Panel B).

**Fig. 2:**
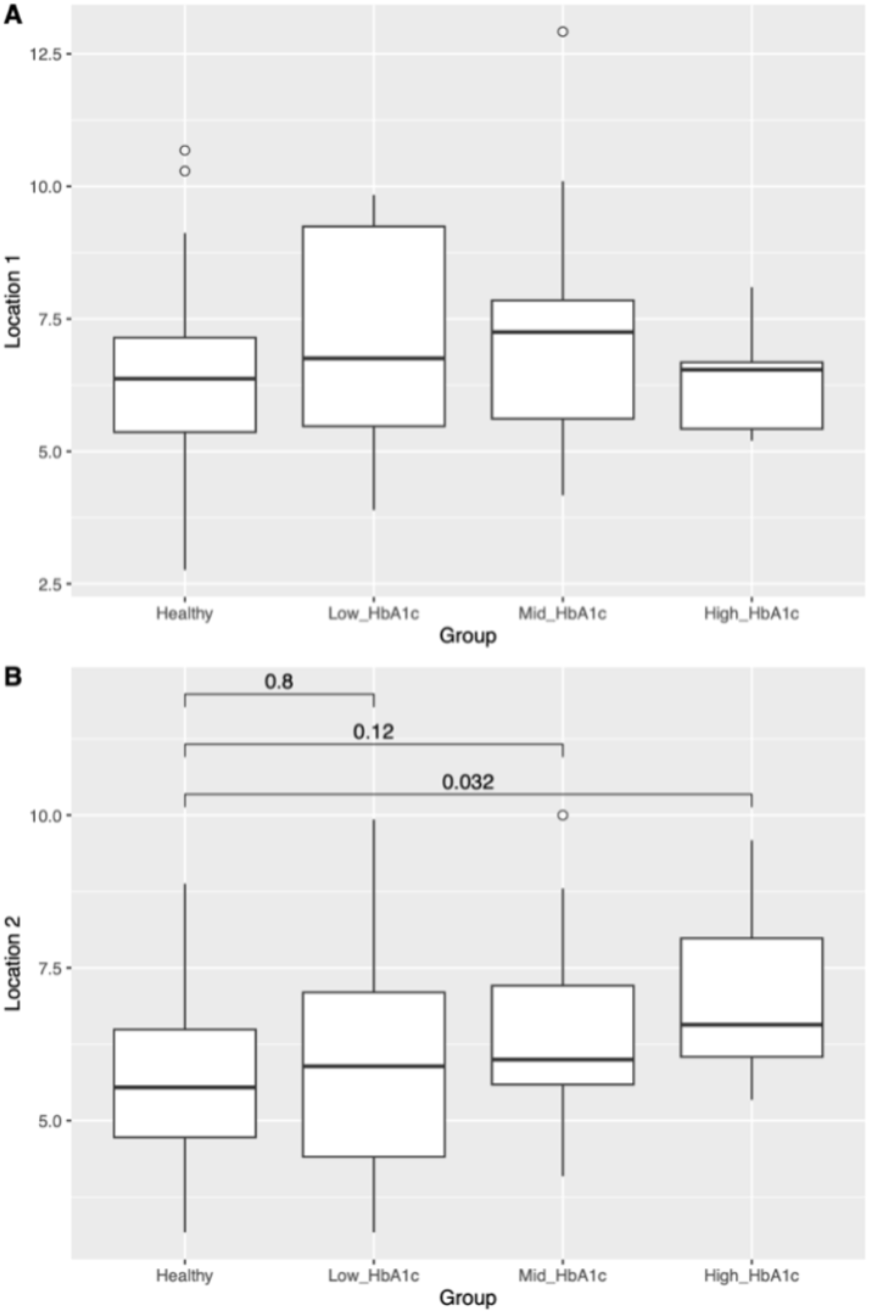
Comparison of absolute CSA between healthy children and children with and HbA1c in the low, mid and high range, respectively, at location 1 (A) and 2 (B), p-values are given above the brackets.

In both the control and diabetes groups, CSA increased with increasing BSA, and the regression coefficient R was bigger in the diabetes groups than in the control group, esp. for the high HbA1c group (Fig. 3).

**Fig. 3:**
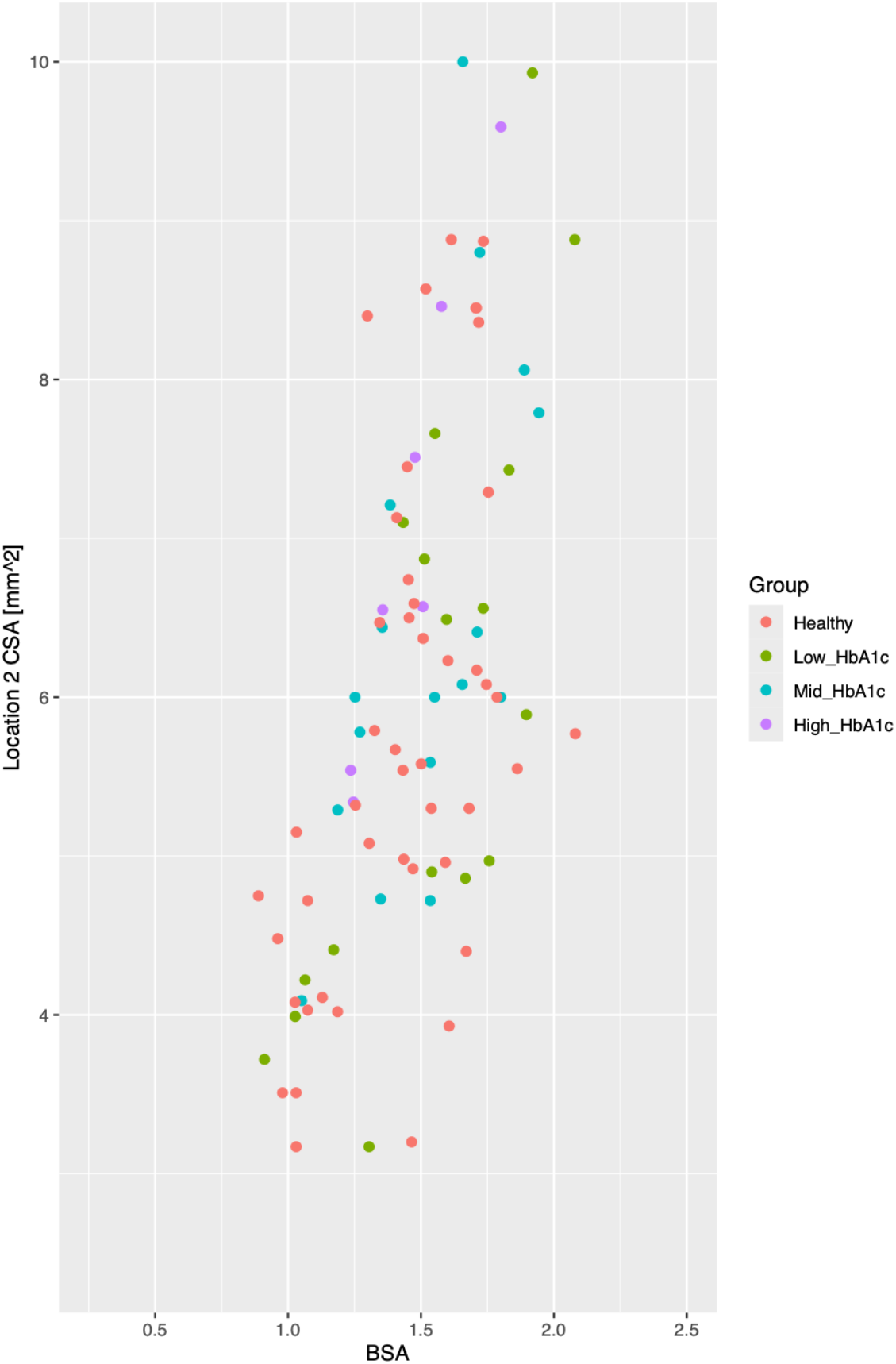
Nerve’s cross-sectional area (CSA) at location 2 in relation to body surface area (BSA) for the same groups as in Fig. 2.

In order to take the correlation of CSA and BSA in children with T1D into account, two approaches were performed. For visualization the ratio between CSA and BSA was calculated and this ratio was plotted against HbA1c for the patients with diabetes (Fig. 4).

**Fig. 4:**
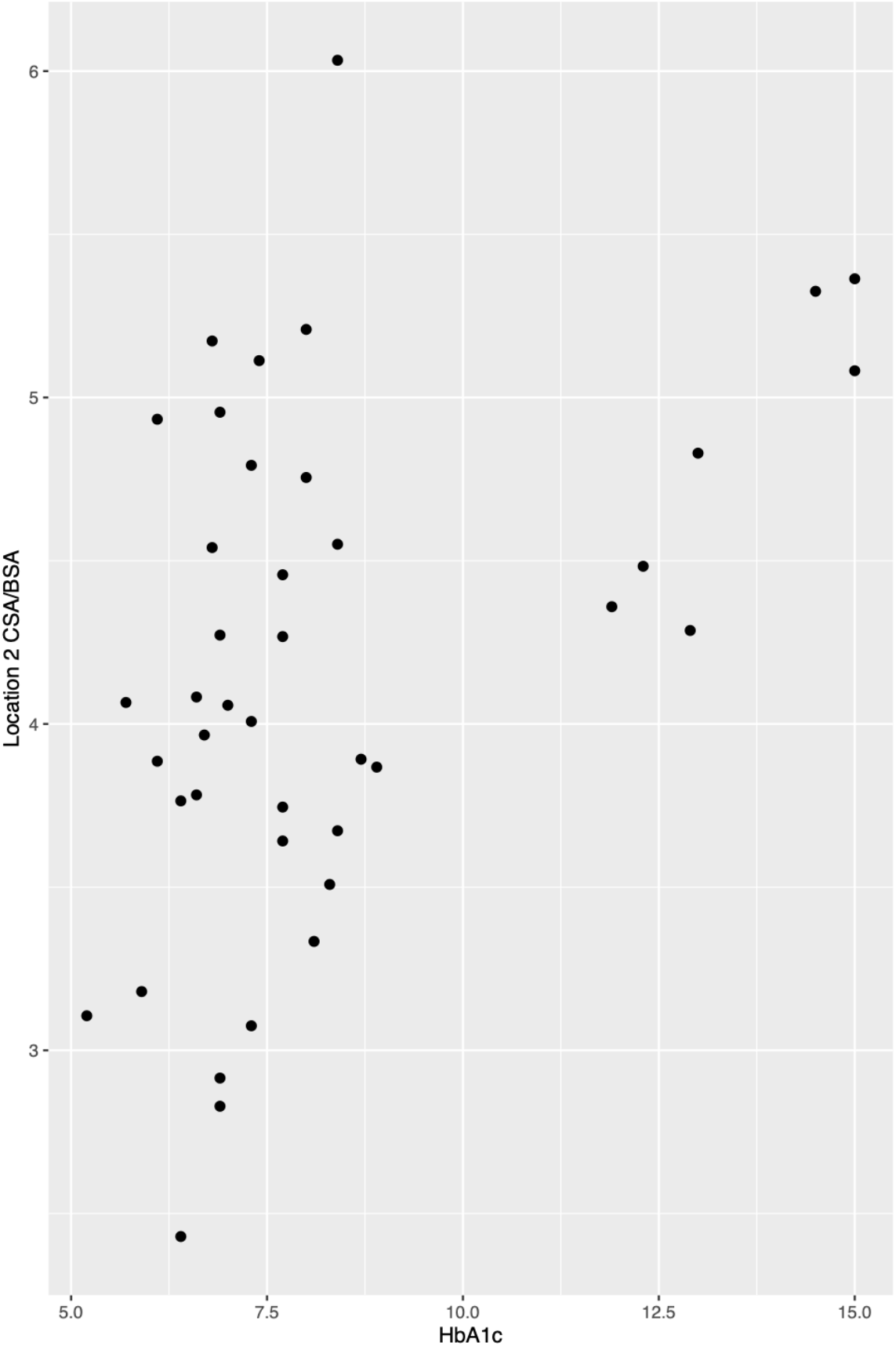
Correlation of the metabolic control and the normalized CSA.

In order to examine whether this finding is statistically significant, a linear model relating CSA to BSA and HbA1c was built (CSA ∼ BSA + HbA1c) and tested for all patients with T1D (Tab. 2). This model showed as expected a highly significant correlation of CSA with BSA, As expected the BSA – reflecting the size of the receptive field of the median nerve – is stronger correlated to the CSA. But more importantly when corrected for BSA using our model the CSA and HbA1c showed a significant and consistent correlation. More precisely the data suggest that a 1% increase in HbA1c increases the CSA of the nerve at location 2 by 0.217 mm^2^ (Tab. 2).

**Table 2:** Multiple linear regression model to explain CSA at location 2 by BSA and HbA1c. Estimate: Estimate of the regression coefficient of the term. CI: Confidence intervall for the estimate.

| Table 2 |  |  |  |
| --- | --- | --- | --- |
| CSA_2 ~ BSA + HbA1c |  |  |  |
| Term | Estimate | 95% CI | p-value |
| BSA | 4.226 | [2.899, 5.554] | <0.001 |
| HbA1c | 0.217 | [0.073, 0.361] | 0.00417 |

## 5. Discussion

The present study highlights the sensitivity of peripheral nerves to damage and the complexity of contributing factors. First, it is demonstrated that, compared to healthy children, the CSA is enlarged in adolescents with insulin-dependent diabetes and HbA1c levels above 9%. This indicates that an insufficient glycemic control engraves CSA enlargement as an expression of poorer nerval health. Second, from an anatomical perspective, this enlargement was only found at location 2, where the nerve is loosely embedded between the deep and superficial finger flexor muscles and hence can expand according to subtle changes in the nerve structure. This is in contrast to location 1, where the nerve is densely surrounded by other uncompressible tissue. Therefore, location 2 can be used as a fiducial marker predicting small fibre neuropathy. Third, given that the CSA critically depends on body size, as represented by the BSA, we controlled for this confounder. The adjusted model showed a strong correlation between glycemic control and nerve thickness at location 2.

One limitation of the study is that nerve ultrasound data are scarce, so we could only design a study that was limited to the median nerve despite the potential existence of other, more suitable nerves. This is especially relevant in the pediatric population where developmental aspects need to be considered, thus larger sample sizes are particularly needed in this population to provide adequate statistical power. The group sizes in this study were too small to test further parameters (e.g., glucose fluctuations).

A strength of this study is that the ultrasound performing doctor was blinded to lab results, esp. HbA1c. This further strengthens our findings. We found a non-invasive diagnostic marker that can be used to detect early nerve damage.

## Data Availability

All data produced in the present study are available upon reasonable request to the authors

## Acknowledgments

The authors thank the team of the neurophysiologic department for their support in conducting the NCS measurements, the secretary of the neuropediatric team for organizing the measurements, and the diabetes team at the Children’s Hospital of Eastern Switzerland for their support. Finally, the authors thank all the children who participated in the study.

## Author Contributions

M.C.E. and M.F. conducted the study, analysis and interpretation of results, writing and finalizing the manuscript. M.C.E. and M.F. contributed equally to this study and are equal first authors.

P.J.B., M.C.E.,M.F. and J.L. were involved in the conception and design of the study. P.J.B., M.F., M.C.E.. and E.W. conducted the study, analysis, and interpretation of results. S.S.O., M.C.E. and U.M. researched patient data and medical records. M.C.E, P.J.B. and all authors edited, reviewed, and approved the final version of the manuscript. M.C.E., P.J.B. and M.F. are the guarantors of this work and, as such, had full access to all the data in the study and take responsibility for the integrity of the data and the accuracy of the data analysis.

All authors have no competing interests to declare.

## Prior Presentations

M.F. Annual Meeting of the Swiss Society of Neuropediatrics, St. Gallen, Switzerland, 11–12 December 2023.

M.C.E. 62nd Annual ESPE Meeting, Liverpool, UK, 16-18 November 2024.

